# Characteristic spatial scales of SARS-CoV-2 pandemics: lessons from mass rapid antigen testing in Slovakia

**DOI:** 10.1101/2020.12.23.20248808

**Authors:** Katarína Boďová, Richard Kollár

## Abstract

Mass antigen testing in Slovakia conducted in October and November 2020 is a source of important data. We perform its statistical analysis and study epidemic geographical patterns. We observe exponentially distributed test positivity and exponential trends in its geographical distribution, and its approximately 10 km spatial characteristic correlation length. A small correlation between positivity in two consecutive testing rounds appeared on the municipalities level but it significantly increased on the counties level. Recent 7-day PCR tests incidence per capita served as a good proxy for antigen test positivity. Positivity of non-residents was higher than of residents when mass testing was offered only in municipalities with the highest positivity in previous rounds. Reduction in positivity in repeated testing increased with the positivity in the earlier round. Our results contribute to better understanding of pandemic data, and aid an assessment of mass testing efficiency, and planning of mitigation measures.

## Introduction

Mass testing is a non-pharmaceutical intervention against SARS-CoV-2 pandemic [1-3]. Antigen tests were used in various mass testing scenarios across Europe in autumn 2020 [4-6] due to their point-of-care application and fast result delivery. These diagnostic tests identify the presence of viral proteins expressed by the SARS-CoV-2 virus. As any mitigation measure antigen tests have their own advantages, limitations and costs to be considered before a mass application [7-11].

The Slovak Republic (population 5.45 million, area 49,035 km^2^) organized mass antigen testing in October and November 2020. It was the first SARS-CoV-2 associated large scale mass testing in Europe. The mass testing was combined with additional mitigation measures to limit mobility, see Supplementary Material S1.4 and [12-14] for the more details. High percentage of public (60%+) tested under similar conditions over a short period of time indicates that test positivity (further referred to as “positivity”) may serve as a systematic measure of the local epidemic situation. The results of the mass testing provide a unique source of information on the mass scale antigen test performance and on emerging geographical epidemic patterns with implications for an efficient and effective selection of pandemic mitigation measures, and particularly for scope and timing of mass testing.

The mass testing campaign in Slovakia consisted of the pilot round (Round 0) organized in a limited area of high incidence, and three consecutive regular testing rounds (Rounds 1-3). Round 1 comprehensively covered the whole country, Round 2 (one week later) was limited to the counties with high Round 1 test positivity and Round 3 (two weeks after Round 2) was limited to municipalities with high positivity in Rounds 1-2.

Round 0 was conducted in 235 testing stations in Bardejov, Dolný Kubín, Námestovo, and Tvrdoš ín counties with the highest current 7-day incidence of positive polymerase chain reaction (PCR) tests on October 23-25, 2020. Three of these countries are located in proximity to the large observed geographical SARS-CoV-2 epidemic cluster in Poland and Czech Republic [15]. In total 140,945 tests were administered, among them 5,594 were positive (3.97%).

Round 1-3 test results in individual municipalities are displayed in Fig.1.

**Figure 1:**
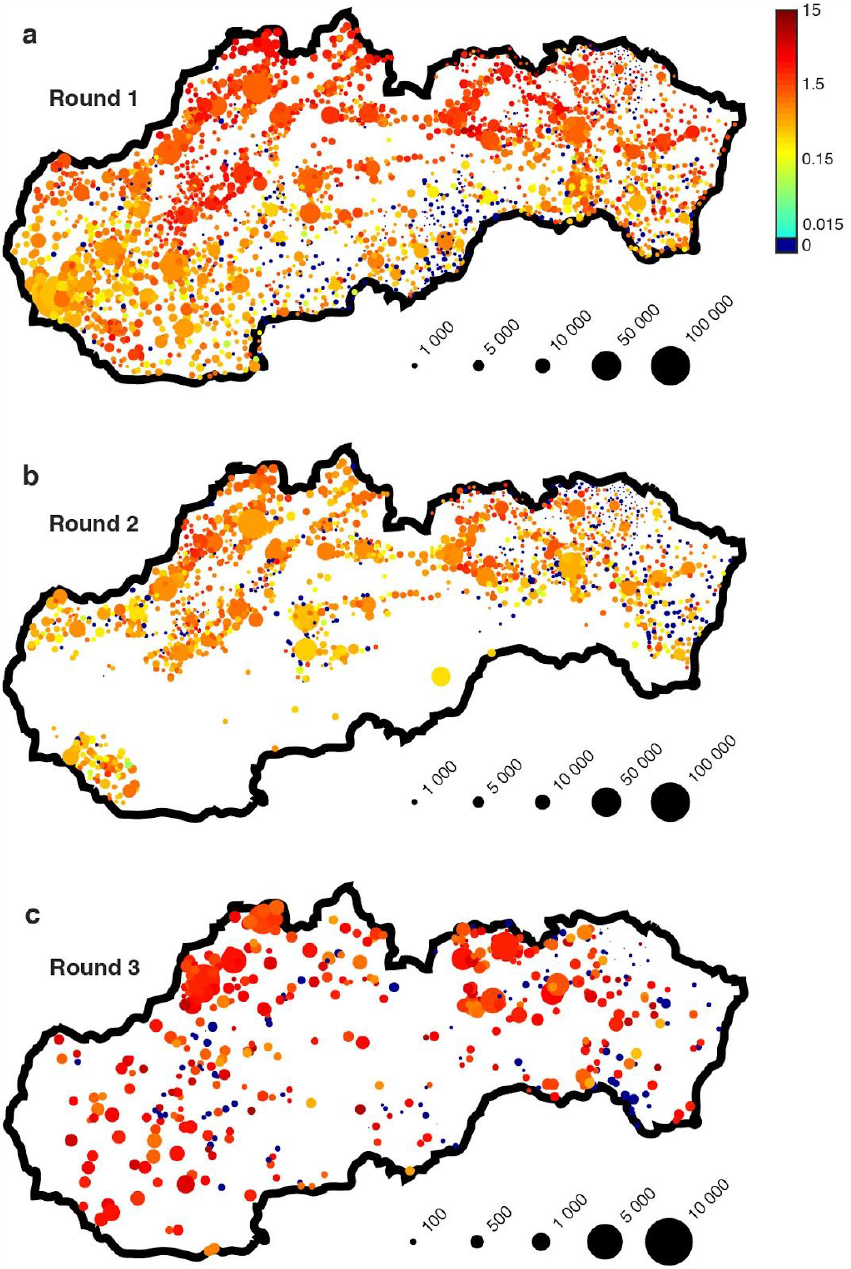
**a-c** Results of mass antigen testing in Rounds 1-3, respectively. Test positivity (%) for individual municipalities is encoded by a logarithmic color scale. The marker size indicates the total number of tests, the size is increased in Round 3 for better data visibility.

Round 1 was conducted in 4782 testing stations (reported on the second testing day) in 2681 municipalities in all 79 counties over the weekend October 31 - November 1, 2020. In total 3,625,332 tests were administered, among them 38,359 were positive (1.06%).

Round 2 was conducted in 2756 testing stations (reported on the second testing day) in 1580 municipalities in 45 counties over the weekend November 8-9, 2020. Testing was restricted to the counties with Round 1 positivity at least 0.7%. In total 2,044,855 tests were administered, among them 13,509 were positive (0.66%).

Round 3 was conducted in 447 municipalities over the weekend November 22-23, 2020. Testing was reduced to the municipalities with Round 1 or 2 positivity at least 1.0%. In total 110,609 tests were administered, among them 2,501 were positive (2.26%). In Rounds 0-2 the untested residents of the municipalities where testing was administered were subject to strict mobility restrictions including inability to go to work for the 7-day or 14-day period following the testing round. Testing in Round 3 was voluntary and no restrictions in addition to general mitigation measures were imposed to untested individuals.

## Methods

Data used throughout this work were published by the Department of Defense and by the Institute for Healthcare Analyses of the Department of Health of the Slovak Republic [16-17]. Round 0 results were reported on the level of the four involved counties without any additional granularity. Round 1-3 results were reported on the level of individual municipalities and city districts. In large cities we combine city districts data into city totals (Bratislava 5 districts, Košice 4 districts) due to a common practice of testing outside of an individual’s household city district. Published data only include the total number of tests performed and the number of positive results without any additional information on demographics and test conditions, except separate reporting of residents and non-residents in each municipality in Round 3.

SD Biosensor Standard Q Covid Ag (SD Biosensor, Inc., Gyeonggi-do, Korea) kit [18], also distributed by Roche [19] was used in all administered tests. It is a rapid lateral flow chromatographic immunoassay for the qualitative detection of specific antigens to SARS-CoV-2 present in human nasopharynx. It provides results within 15-30 minutes that are evaluated by naked-eye. The manufacturer claims sensitivity 96.52% (95% CI 91.33-99.04%) and specificity 99.68% (95% CI 98.22 – 99.99%) in the test package leaflet [18]. See Supplementary Material S1.3 for a further survey of 10 validation studies [20-29] with sensitivity median 78.1% and average 78.9%, and specificity median 99.4% and average 98.1%. The sample tested in the mass testing in Slovakia may significantly statistically differ from the validation studies samples. Dependence of antigen test sensitivity on viral load was studied in [30]. The lower specificity bound (99.6%) in mass testing in Slovakia was estimated in [31]. Test methodology and setup are described in detail in [12], see also additional information in Supplementary Material S1.4.

All data within this work were fitted with built-in tools for nonlinear regression (`nlinfit` and `nlparci`) and a Curve Fitting Toolbox for weighted linear regression in MATLAB©. The full code generating the figures in this work along with all used data are provided in the Supplementary Material S2.

Global Moran’s I statistics measuring spatial autocorrelation is used for the analysis of geographical patterns of positivity in Round 1. It provides statistics for spatially irregular data confined in an irregular geographical region with an uneven density of locations in space. The value of the Moran’s I statistics is computed for a particular choice of spatial weights that encode contiguity of regions and their mutual influence as a function of their distance:

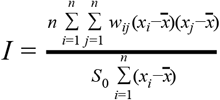

where x_i_ is the positivity in region i, n is the total number of regions, 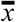 is the average test positivity in all regions, and S_0_ is a sum the spatial weights w_*i,j*_ : 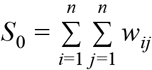.

To identify the characteristic length of test positivity we introduce square tilings of Slovakia with the increasing dimension of a tile side (3, 4, …, 50 km). Only tiles overlapping with the area of the Slovak Republic are included. For each tile dimension we consider three alternative tile contiguity definitions: two tiles are contiguous if their centers are no more than 1-, 1.5-, and 4-times the tile dimension apart. The maximum number of neighbours of a tile is 4, 8, and 48, respectively, see Fig 3a. All tiles that are contiguous contribute equally to the Moran’s I statistics weight function, non-contiguous tiles have zero weight.

To compute significance levels for the global Moran’s I we use random shuffling of the tile positivity values while keeping their locations unchanged. We generate 4999 shufflings to obtain a null distribution of Moran’s I under assumption of spatial independence.

To analyze the geographical distribution of Round 1 positivity we average the data over a rectangular grid (2 x 3 km) covering the area of Slovak Republic (see Fig. 4a). Local positivity in each grid rectangle is calculated as positivity of all tests administered in the municipalities centered within the rectangle. For visualization purpose only, positivity on the rectangular grid in Fig. 4a is smoothened by diffusion (diffusion coefficient 0.05 km^2^ /s, final time 0.45 s) to effectively average out the high level of local variation.

## Results

A statistical summarization of mass testing results is presented in Fig. 2. In Rounds 1-2 positivity in individual municipalities is well approximated by an exponential distribution (see Fig. 2a; note the log-linear axes scale). Round 3 positivity of tests of residents is also well approximated by an exponential distribution. The same trend is not observed for all tests in Round 3 (not shown) as the results were skewed by high participation of non-residents (22.6% of all tested samples) and their high average positivity (3.88% compared to average residents positivity 1.13%). See Fig. 2b for the graphical comparison of positivity distribution of residents and non-residents in Round 3 in individual municipalities.

**Figure 2:**
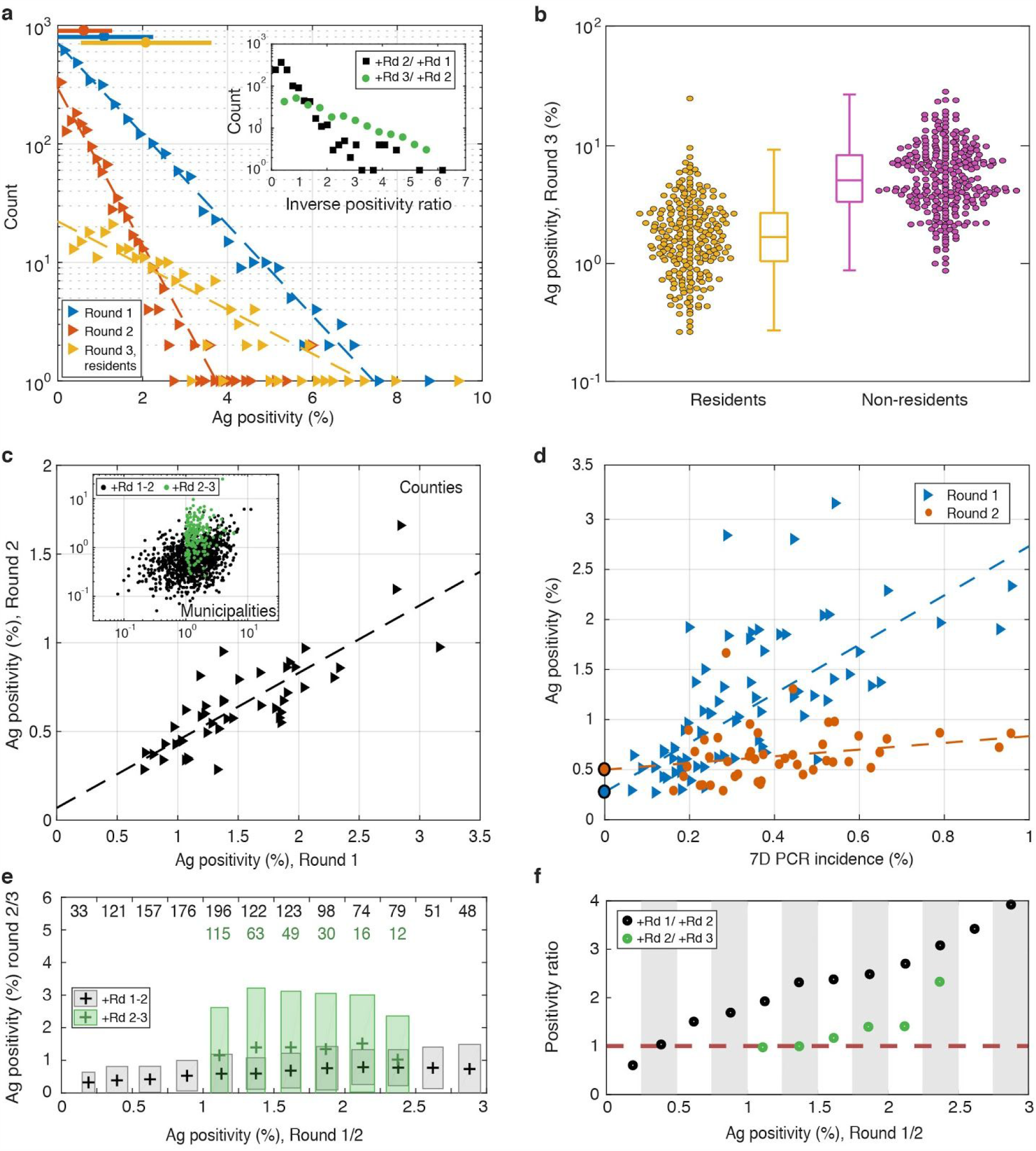
Statistical data analysis. **a** Histograms of positivity in municipalities in Rounds 1-3. Horizontal color bars on the top indicate mean±std (Round 1: n=2681, Round 2: n=1580, Round 3: n=447). Each triangle represents a number of municipalities within the corresponding positivity bin. Dashed lines correspond to exponential fits. Inset shows distributions of the inverse relative change of positivity between Rounds 1-2 and between Round 2 and residents in Round 3. **b** Residents (n=233) vs. non-residents (n=265) positivity in Round 3. Zero data is not displayed. The central mark shows the median, the box shows the 25th and 75th percentiles, the whiskers show the most extreme values not considered as outliers. **c** Positivity in Round 2 vs. Round 1. Data for counties (n=45) in Rounds 1-2 are shown in the main plot, data for municipalities with positive antigen results are shown in the inset (Rounds 1-2: n=1098, Rounds 2 and Round 3 (residents): n=186). The linear fit shown has R-squared 0.66 (p<0.001 for the slope estimate). Only data from locations with both compared testing rounds are shown. **d** Positivity in Rounds 1-2 against 7-day PCR incidence prior to Round 1 rescaled by population size. The linear fits shown have R-squared 0.46 (Round 1) and 0.06 (Round 2). **e** Comparison of positivity in Round 2 vs. Round 1 and Round 3 (residents) vs. Round 2. Data on the horizontal axis are binned into bins of width 0.25 (%), the color box in each bin indicated mean±std interval for both variables (n shown on the top). **f** The ratio of means of positivity in two consecutive rounds, bins with width 0.25% in the earlier round; based on **e**.

The scale S and the rate r parameters of the exponential form y=S*exp(-r*x) fit to the data with 95% confidence intervals and the coefficient of determination R-squared are Round 1 (n=2681): R-sq = 0.9975, S = 706.17 (692.61, 720.00), r = 0.88 (0.86,0.91), Round 2 (n=1580): R-sq = 0.9185, S = 293.94 (263.49, 327.90), r = 1.52 (1.29,1.76), Round 3 (n=447): R-sq = 0.8517, S = 22.23 (19.23, 25.71), r = 0.43 (0.35,0.51).

A similar pattern is observed in the distribution of the inverse ratio of positivity in individual municipalities between Rounds 1 and 2, and between Rounds 2 and 3 (see Fig 2a inset, log-linear scale).

A comparison of positivity in two consecutive rounds is displayed in Fig. 2c. Small correlation is observed on municipalities level (see Fig. 2c inset), Rounds 1 and 2 positivity correlation R-squared value is 0.10 (n=1580), the slope 0.17 (p<0.001, 95% CI 0.15-0.20). Rounds 2 and 3 (residents) positivity correlation R-squared is 0.02 (n=314). Increased correlation in test positivity appears if data are weighted with the total volume of tests in the earlier of the two compared rounds: R-squared value for a weighted linear correlation of positivity in Rounds 1 and 2 is 0.23 (n=1580), the slope 0.24, (95% CI 0.22-0.28). R-squared for a weighted linear correlation of positivity in Rounds 2 and 3 (residents) is 0.01 (n=314).

Significantly higher correlation is observed on the counties level (Fig. 2c main panel). Rounds 1 and 2 positivity correlation R-squared is 0.66 (n=45), the slope 0.38 (p<0.001, 95% CI 0.30-0.46), the absolute coefficient 0.07 (p=0.31, 95% CI -0.07-0.21). The linear correlation weighted by the total number of tests in the earlier round has R-squared 0.65 (n=45), the slope 0.34 (95% CI 0.27-0.42), and the absolute coefficient 0.12 (95% CI 0.00-0.25).

A comparison of positivity (on municipalities level) in pairs of consecutive rounds is shown in Fig. 2e. Results of Rounds 1-2 and Rounds 2 and 3 (residents) are compared. Data in the earlier of the two compared rounds are binned to smooth out an individual variation of positivity: municipalities with positivity 0.00-0.25%, 0.25-0.50%, up to 2.75-3.00% are clustered together. Two-dimensional means and confidence intervals are displayed indicating the mean ± standard deviation of the positivity in the earlier and the later of the two compared rounds.

Figure 2f shows the ratio of means of positivity in the earlier and in the later of the two compared rounds displayed in Fig. 2e (bins with less than three data points are omitted). The ratio of mean positivity in Rounds 1 and 2 increases with the positivity in Round 1 for the bins up to 3%, and it is approximately twice as large as the mean positivity ratio in Rounds 2 and 3. Moreover, the mean ratio factor between Rounds 2-3 does not change significantly for the bins with positivity between 1% and 2%. The ratio of the mean positivity in Rounds 1 and 2 for a bin with Round 1positivity <0.5% does not exceed the value 1 indicating an increase of positivity in Round 2 compared to Round 1. The ratio of means of positivity is approximately 1 between Rounds 2 and 3 (residents) for bins with Round 2positivity <1.5%.

The relationship between antigen tests positivity and PCR test incidence in 70 individual counties and two cities (Bratislava and Košice) is displayed in Fig. 2d (PCR incidence is only reported on counties level). Antigen tests positivity in Rounds 1 and 2 is compared with the 7-day incidence of positive PCR tests divided by the county population one day prior to the start of Round 1 (the total of positive PCR tests, Oct 24-30). It is the last PCR data point before the start of the mass testing campaign. Unavailability of testing stations, low numbers of indications for testing by Regional Public Health Authorities, etc. caused disruptions in PCR data after mass testing commenced. Round 2 mass testing data are thus compared with the 7 days delayed PCR incidence data.

The best weighted linear fit of the form y = ax + b to Rounds 1-2 data is shown in Fig. 2d. The weight of each data point corresponds to the total number of antigen tests administered in the county. The inferred parameter values for Round 1 are a=2.46 (95% CI: 1.82-3.10), b=0.28 (95% CI: 0.04-0.51), R-squared 0.46 (n=72). For Round 2 the parameters are a=0.34 (95%CI: -0.07-0.74), b=0.50 (95% CI: 0.32-0.68), R-squared 0.06 (n=45).

Additional information is provided by the weighted linear regression to the reduced dataset of the counties with small antigen test positivity (< 1%) in the corresponding mass testing round (not shown). R-squared values are 0.11 (n=34) and 0.15 (n=43) for Rounds 1 and 2, respectively.

Quantitative information about the geographical correlation of Round 1 positivity is encoded by the global Moran’s I statistics (see Methods) displayed in Figure 3. At small distances (< 5 km) low positive spatial autocorrelation is observed, reflecting high variability in test positivity between individual (mostly small) municipalities, see Fig. 3a. At large distances (> 20 km) a steady decay of spatial correlation is observed reflecting a low level of interconnection between more distant regions. Maximum value of the autocorrelation statistics is reached for 10 km consistently for three different neighboring regions definitions (see Methods). The weight function representing the neighboring region relation influences only an overall magnitude of the test statistics but not the maximum location. The characteristic correlation length scale of Round 1 positivity is therefore approximately 10 km.

To confirm the statistical significance of the computed global Moran’s I Fig. 3b shows the distributions of Moran’s I under the null hypothesis of no spatial correlation of data for the characteristic correlation length (tile size 10 km) for all three contiguity definitions. In all three cases the maximum value of Moran’s I did not exceed 0.1 for 4999 random data permutations, compared to the values approximately 0.69, 0.66, and 0.49 for unpermuted data.

**Figure 3:**
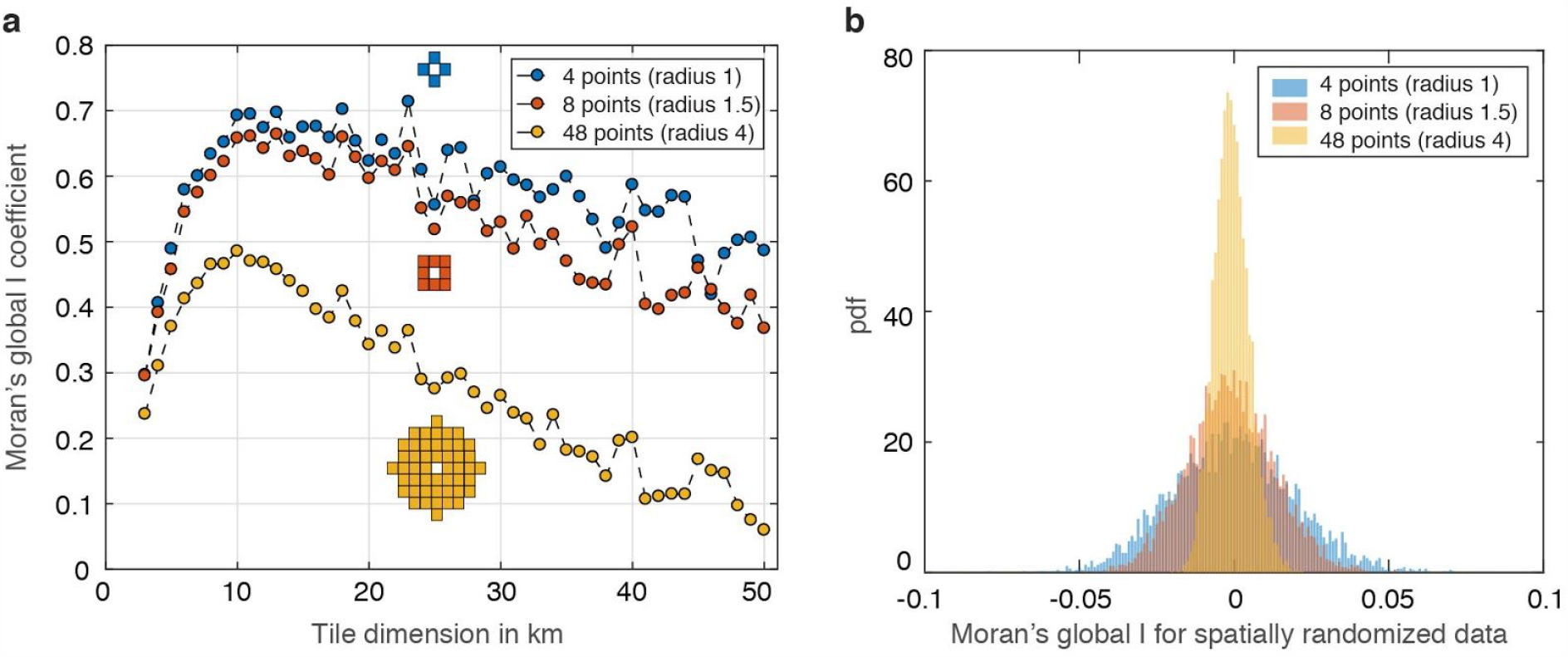
Spatial correlation of Round 1 positivity. **a** Moran’s I statistics plotted against the dimension of a square tile covering the area of Slovakia. For each tile dimension Moran’s I values for three alternative weight functions are displayed, see Methods for more details. The peak at 10 km indicates the characteristic correlation length scale. The value of Moran’s I at 10 km for the three choices of weight matrices is 0.6926 (blue), 0.6581 (red), 0.4854 (yellow), all with p<0.001 (n=1323 square tiles with nonzero positivity). **b** Distribution of global Moran’s I for randomly reshuffled data (4999 replicates) for the three weight functions and tile dimension 10 km. All values obtained by reshuffling were smaller than 0.1 implying p-values < 0.001.

Geographical patterns in Round 1 positivity are demonstrated by spatial gradients displayed in Fig. 4. A south-west to north-east gradient is apparent in Fig. 4a. Its quantification in the observed data is blurred by a convolution of signals from all directions, and particularly from multiple dominating sources located in areas of high test positivity.

**Figure 4:**
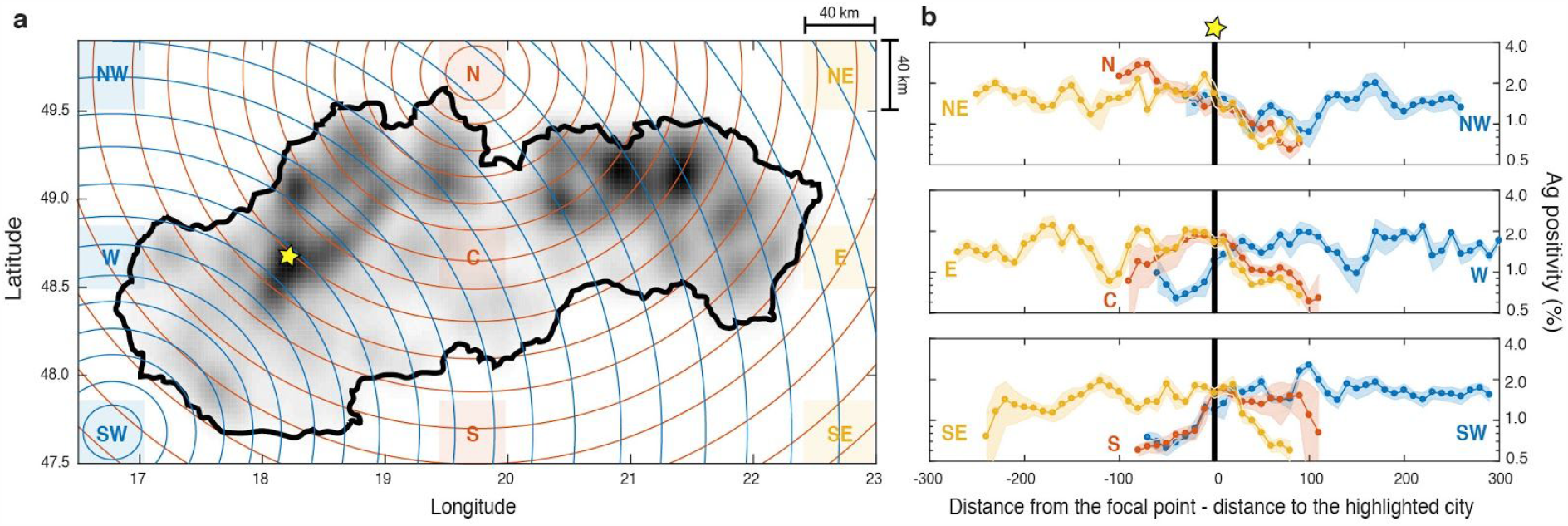
Geographical distribution of positivity. **a** Round 1 positivity (increasing light to dark gray) on a fine rectangular grid is for visualization purposes smoothened by diffusion (see Methods). **b** Dependence of average positivity (log scale) on the distance from 9 base areas. See **a** for the base areas locations (40 km x 40 km each). 247 regularly spaced grid points are selected in each region. For each grid point the average test positivity in individual municipalities located within annular regions 10 km-wide centered at the grid point (see **a** for an illustration for two particular grid points in SW and N base areas) is calculated. The curves in **b** display the average positivity (over all grid points in the same base area) at each distance with shaded mean±std. Distance from each grid point in the averaged ensemble in each base area is normalized so that the reference point of high positivity (indicated by a yellow star in **a**) is shifted to the origin on the horizontal axis (distance 0).

The convolution effects are partially eliminated by spatial averaging of positivity observed from distant locations. Nine base areas (denoted as N, NE, E, SE, S, SW, W, NW, W, and C) are located near the map edges in the cardinal and the intercardinal directions and at the map center. These base areas are located in Poland, Ukraine, Hungary, Austria, Czech Republic and Slovakia. The dependence of Round 1 positivity on the distance from all 9 individual base areas together with the 68% confidence interval (mean±std) is displayed in Fig. 4b. The distance 0 corresponds in all plots to the distance from each base area to the high positivity area marked by a yellow star in Fig. 4a.

Trends in positivity emerge locally in Fig. 4b. On a log-linear scale they correspond to spatial exponential dependence of positivity on the distance. A good overall agreement with the exponential trend is observed particularly from the base area N. Beyond a small increase of positivity at short distances positivity systematically decreases approximately exponentially with the increasing distance. The opposite spatial exponentially increasing trend is observed from the base areas SW and S up to the distance at which the annular regions reach the clusters of municipalities of high positivity. Beyond this distance positivity reaches a plateau.

## Conclusions and Discussion

Mass antigen testing in Slovakia generated the first publicly available dataset that comprehensively characterizes the local epidemiological situation during the SARS-CoV-2 pandemics on the level of individual municipalities across the whole European country. The data provide information on antigen test performance on a large scale. Under the assumption that the test positivity quantitatively represents the local epidemiological situation the data also contain information on epidemic geographical spreading patterns. The data indicate the importance of a spatial scale in data analysis, as individual municipalities data and data averaged on a very fine scale may eventually conceal important trends.

One of the central questions is the amount of reduction of infected individuals by repeated mass testing. Our data analysis offers only indirect information on this subject in the quantification of the ratio of positive antigen tests in two consecutive rounds. On one hand, the combination of two consecutive testing rounds 7 days apart with mandatory strict mobility reduction for untested individuals yielded on average an increase of positivity in the later round in the municipalities with low test positivity in the earlier round (<0.25%). The increase may be caused by statistical randomness in the tested sample and therefore this phenomenon needs to be studied further. On the other hand, in municipalities with higher test positivity in the earlier round (>0.5%) the test positivity in the later round was reduced on average. The increasing trend in test positivity ratio between Rounds 1 and 2 with respect to the Round1 positivity indicates that the repeated mass testing is the most effective epidemic reduction measure in regions with the highest positivity.

Round 1 mass antigen tests positivity is correlated with the most recent 7-day PCR positive tests incidence per capita on the counties level. This suggests PCR tests data provide a good proxy for the epidemiological situation under an assumption that the PCR testing capacity is sufficient. However, PCR incidence data delayed by one week did not provide significant information about the Round 2 positivity.

Participation in mass testing decreased significantly when testing in the later round was not connected with the mobility restrictions of untested individuals and the mass testing was only administered in municipalities with the high test positivity in the previous rounds. An average Round 3 positivity was systematically higher than in the earlier rounds indicating that the testing without mandated mobility restrictions for untested attracts a smaller testing sample but at the same time a sample with a higher test positivity. Moreover, free voluntary mass antigen testing that was offered only in certain locations in Round 3 showed that the non-residents who participated in testing had higher averaged positivity than residents. That is despite the fact that testing was organized in the municipalities with the highest positivity in the previous rounds. These results indicate that the free testing may attract under these conditions a population sample with high positivity and such an effort may provide an efficient tool to identify antigen test positive individuals.

Geographical spreading is an important pandemic aspect [32-35]. The data from mass testing in Slovakia reveal spatial patterns and characteristic scales. The test positivity on the municipality level is exponentially distributed for both testing with and without mandatory mobility restrictions for untested individuals. Our further data analysis reveals that the distribution is connected with the exponential patterns in spatial spreading. In Slovakia it is demonstrated by observed exponential increase of averaged test positivity in certain geographical directions. Trends in geographical spreading, particularly if observed from areas in other countries, as in Fig. 3a can be useful in pandemic mitigation strategies. They may serve as an informed estimate for the decisions on border closing, mobility mitigation measures, and other interventions.

Traditional epidemiological models predict approximately exponential temporal growth of a fraction of infected in a population in the early stages of epidemic [36]. A simple mechanism of a constant speed radial epidemic initiation and spreading from the localized sources is therefore consistent with spatially exponential trends observed in our analysis. This is in agreement with the model in [37] that provides an explanation for the algebraic growth trends observed in epidemiological data globally [38,39].

Finally, we also identify the characteristic spatial correlation length of test positivity of approximately 10 km, i.e., the size of natural square regions that show the highest level of spatial autocorrelation of test positivity with their neighbours is 10 km x 10 km. The value is inferred from the Global Moran’s I statistics used in spatial studies of pandemics in China [32-35]. The level of spatial correlation of positivity in mass testing in Slovakia was significantly higher than observed in [32,33]. The same statistics for the total number of tests administered in Round 1 reveals different trends (see Fig. S1). This indicates that the observed characteristic length scale of test positivity cannot be directly explained by the test volume that serves as a proxy for the total population. The characteristic test positivity correlation length may aid an efficient localization of mitigation measures as it provides a quantitative estimate of the size of an area with the highest probability of correlation in the vicinity of known areas of high infection incidence.

The data and its analysis discussed here have systematic limitations. All findings are limited in scope to the epidemiological situation in Slovakia during October-November, and any inference from the data applied to other settings needs to be taken with precaution. The same is true about an inference of infection prevalence from the positivity data as no direct relation between these two factors was established yet.

Systematic limitations stem from test parameters, testing procedures and test evaluations, see Supplementary material S1.4 for more details. Up to this date there is a lack of scientific validation of the test parameters used on a general population sample dominated by asymptomatic individuals. No test validation was conducted during the mass testing in Slovakia, test results were not confirmed by other laboratory diagnostics, test subjects were often not at room temperature prior to sample collection, the sample collection was performed by volunteer medical personnel that did not receive proper training how to perform the pharangonasal swabs, no specialized training was provided for test evaluation, 20 Euro risk compensation was paid to staff collecting the sample for each positive test.

Test parameters (sensitivity and specificity) may be sample dependent as their sensitivity depends on distribution of viral loads in the sample [30]. Areas with fast epidemic growth that are dominated by fresh infections have a higher apparent average viral load in routine PCR testing observed in the data than areas without such growth or on an epidemic decline [40]. Any eventual validation study should therefore be conducted under various epidemiological conditions.

The individual municipality data in Rounds 1 and 2 may be influenced by testing of individuals at testing stations outside of their area of the residence. The data may also contain duplicities caused by individuals tested repeatedly within one round. The relation between test positivity in individual rounds is also influenced by the local speed of epidemic growth. The local effective reproduction number may significantly contribute to the relative change of the test positivity between two rounds. This factor prohibits a simple interpretation of the reduction of the infected in the population by the mass testing effort.

The results of the mass rapid antigen testing in Slovakia were also studied in [12,31] (on the county-level data). There are significant differences between the results presented here and in [12] that does not analyze Round 3 data. Within this work we refer to test positivity and do not relate it directly to infection prevalence due to the uncertainty in test parameters on a population wide sample under various epidemiological settings. We do not attempt to evaluate the success of the testing campaign as the long term effects of the campaign are not yet analyzed and Slovakia recorded increased mobility trends immediately after the mass testing campaign and faced a surge of infections 3 weeks later. Unlike [12] our analysis reveals a large degree of geographical heterogeneity primarily demonstrated by the exponential distribution of the test positivity on municipalities level and also by exponential trends in geographical spreading. Although we observe a systematic reduction of positivity in repeated testing on the counties level we detect an increase of test positivity in repeated testing in low positivity municipalities (<0.25%).

The exponential distribution of test positivity observed in municipalities data in Rounds 1-3 is in contrast with the study [41] where the observed incidence distribution across individual countries followed a power-law distribution, see also Fig. S2. The type of distribution and its decay rates indicate that there are disproportionate numbers of municipalities with extreme test positivity values, only a few with relatively high positivity and many with relatively low positivity. Such an information may aid resource allocation during the pandemic and planning of mitigation strategies.

## Data Availability

Analysis is based on publicly available data

## Acknowledgements

This work has been supported by the Slovak Research and Development Agency under the Contract Nos. APVV-18-0308 (RK) PP-COVID-20-0017 and by the Scientific Grant Agency of the Slovak Republic under the Grants Nos. 1/0755/19 and 1/0521/20.

## Supplementary Material S1

### S1.1 Moran’s global I statistics for the total number of administered tests

**Figure S1:**
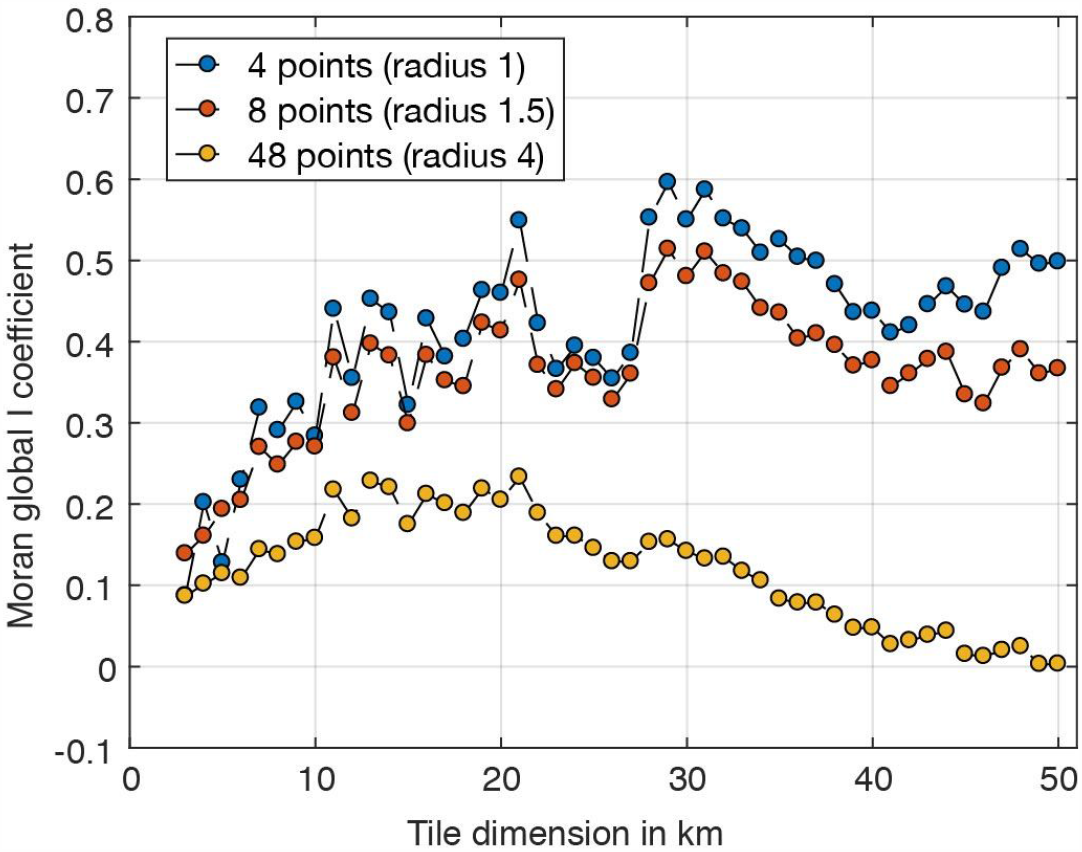
Spatial autocorrelation of the number of tests performed in Round 1 measured by the global Moran’s I statistics. We consider square tilings of Slovakia with increasing dimensions of the tile side (3, 4, …, 50 km), on the horizontal axis. For each tile dimension we use three alternative spatial weight functions, defining contiguity when distance between the tile centers is no more than 1-, 1.5-, and 4-times the tile dimension (blue, red, and yellow, respectively). All tiles that are contiguous contribute to the Moran’s I statistics with the same weight, while non-contiguous tiles have weight 0. The code in MATLAB© is provided in the Supplementary Material S2.

### S1.2 Global test positivity distribution

**Figure S2:**
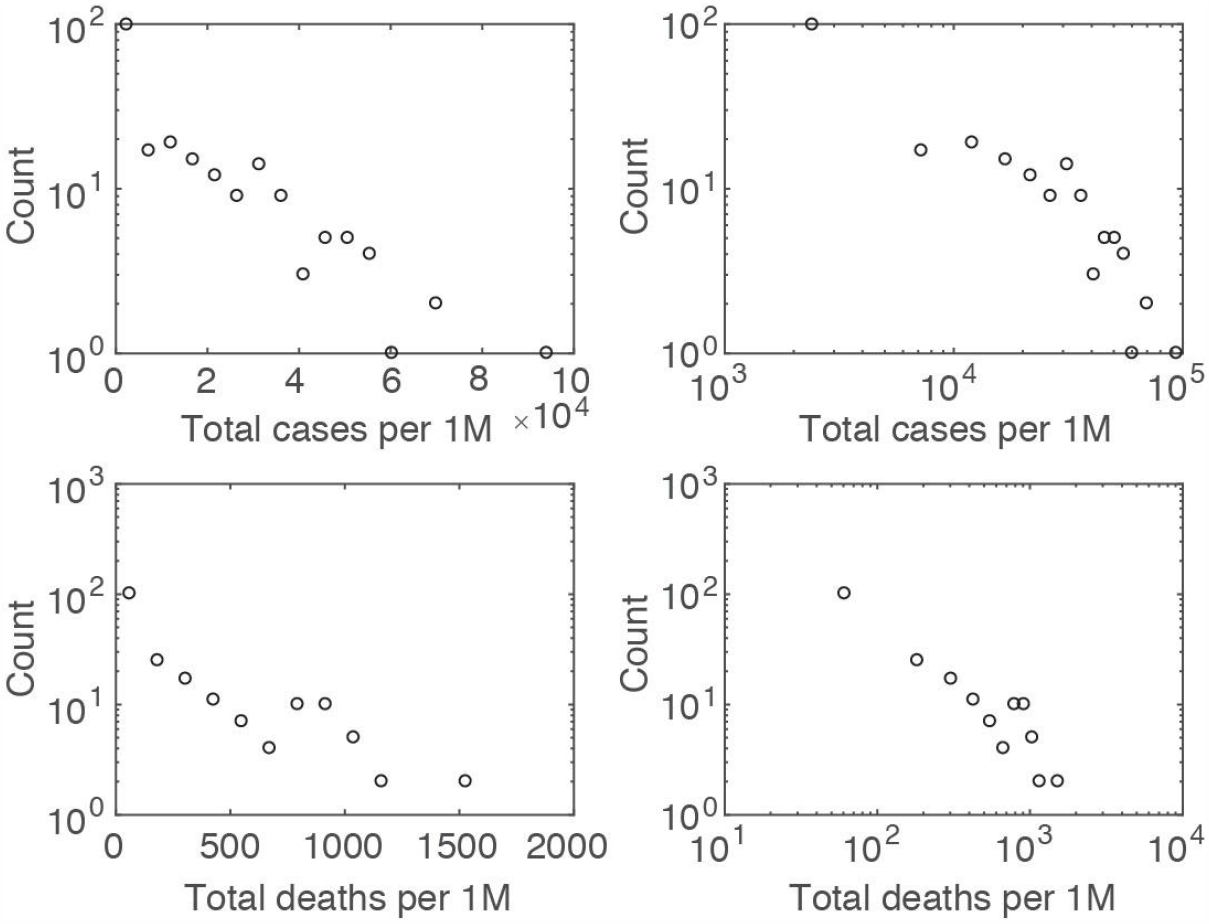
Histogram of total cases and total deaths per one million people reported in individual countries around the world on December 2020 (source: worldometers.info) on a log-linear (left) and log-log (right) scales (n=215 for the total cases, n=194 for the total deaths). A truncated power-law distribution of total cases was observed in [1] in March 2020, in the early phase of the pandemic, when the virus was newly observed in most of the studied countries. No consistent exponential or power-law trends are observed in the current data, except the exponential trend in total deaths for countries between 100 - 600 total deaths per one million people.

### S1.3 Validation studies of the SD Biosensor Standard Q Covid Ag test

**Table S1:** Validation studies of the SD Biosensor Standard Q Covid Ag (SD Biosensor, Inc., Gyeonggi-do, Korea) test [12] also distributed by Roche [13]. *Test #2 in the validation study is SD Biosensor Standard Q Covid Ag test.

| Source | Country | Sample Size | RT-PCR positive % (n) | Sensitivity % (n) | Specificity % (n) |
| --- | --- | --- | --- | --- | --- |
| Berger A, et al. [2] | Switzerland | 529 | 3.6% (191/529) | 89.0% (170/191) | 99.7% (1/338) |
| Linder AK, et al. [3] | Germany | 289 | 13.5% (39/289) | 79.5% (31/39) | 99.6% (1/250) |
| Krueger LJ, et al. [4] | Germany/UK | 1263 | 3.7% (47/1263) | 76.6% (36/47) | 99.3% (9/1216) |
| Cerutti F, et al. [5] | Italy | 330 | 33.0% (109/330) | 70.6% (77/109) | 100.0% (0/221) |
| Sahar MK, et al. [6] | Egypt | 100 | 80.0% (80/100) | 68.7% (55/80) | 95.0% (1/20) |
| FN Motol [7]* | Czech Republic | 591 | 37.6% (222/591) | 62.6% (139/222) | 99.5% (2/369) |
| FIND [8] | Brazil | 400 | 26.5% (106/400) | 88.7% (/106) | 97.6% (7/294) |
| Nalumansi A, et al. [9] | Uganda | 262 | 34.3% (90/262) | 70.0% (63/90) | 92.4% (13/172) |
| Chaimayo C, et al. [10] | Thailand | 454 | 13.2% (60/454) | 98.3% (59/60) | 98.7% (5/394) |
| Iglói Z, et al. [11] | Netherlands | 970 | 19.2% (186/970) | 84.9% (158/186) | 99.6% (3/784) |
| Average |  |  |  | 78.9% | 98.1% |
| Median |  |  |  | 78.1% | 99.4% |

### S1.4 Additional mitigation measures imposed during mass antigen testing in Slovakia and specifics and limitations of tests and testing procedure

All individuals with positive test results were ordered to isolate for 10 days with their whole households. All persons with negative test results in the pilot, and in Rounds 1-2 of mass testing were issued an official (blue) certificate that allowed them to avoid some of the strict measures enforced during the following 7-day period (or 14-day in counties without the second testing round). During the period all persons without a confirmation of a negative antigen test or a recent negative polymerase chain reaction (PCR) test were subject to a strict lockdown. They were allowed to leave their household only during the night (01:00-05:00 am), or to leave their household to take a PCR or an antigen test, to visit the nearest grocery store or a pharmacy, or to get medical care. They were also allowed to provide assistance and a personal care for their close persons or livestock, to walk pets up to 100 meters away from their household, and to attend a funeral. Those with negative tests were in addition allowed to travel to work, accompany their children to school, to visit post offices, insurance companies, drycleaning, car repair shops, and petrol stations. They were allowed to spend time in nature within their counties outside of urban areas.

More details on extent and timing of mitigation measures during testing and testing procedures can be found in [14].

According to the test package leaflet “the test is for administration by healthcare workers and labs only, as an aid to early diagnosis of SARS-CoV-2 infection in patients with clinical symptoms with SARS-CoV-2 infection. It provides only an initial screening test result … The result of this test should not be the sole basis for the diagnosis; confirmatory testing is required” [12]. During the mass testing in Slovakia test results were not confirmed by other laboratory diagnostics and most of the tested individuals were asymptomatic.

The manufacturer also recommends that the tess are at the room temperature prior to sample collection. However, during the mass testing, a large number of testing stations were for epidemiological reasons located in outdoor settings and the temperature at many testing stations was under the CDC recommended room temperature (15-30°C) before use [15].

The sample collection was performed by volunteer medical personnel that did not receive proper training how to perform the pharangonasal swabs. Also no specialized training was provided for an evaluation of the tests. The bias in data may be caused by 20 Euro risk compensation to staff collecting the sample for each positive test, although the test was evaluated by a different member of the testing team that did not receive any risk compensation.

Tests were administered at temporary set testing stations. They were staffed by volunteers, each testing team included among others two medical professionals who collected samples and evaluated tests and one member of the army forces who coordinated testing locally and reported the data to the army regional headquarters for data collection. No specific training was provided to medical staff. Testing stations were located both indoors and outdoors, a minority of testing stations were drive-through.

